# Machine Learning-Based Identification of High-Risk Patterns in Atrial Fibrillation Ablation Outcomes

**DOI:** 10.1101/2024.11.27.24318097

**Authors:** Mustapha Oloko-Oba, Yijun Liu, Kathryn Wood, Michael S. Lloyd, Joyce C. Ho, Vicki Stover Hertzberg

## Abstract

**Background:** Atrial fibrillation (AF) is one of the most common types of cardiac arrhythmias, often leading to serious health issues such as stroke, heart failure, and higher mortality rates. Its global impact is rising due to aging populations and growing comorbidities, creating an urgent need for more effective treatment methods. AF ablation, a key treatment option, has success rates that vary widely among patients. Conventional predictors of ablation outcomes, which primarily rely on sociodemographic and clinical factors, fall short of capturing the heterogeneity within patient populations, highlighting the potential for data-driven methods to provide deeper insights into procedural success.

**Objectives:** To uncover meaningful patient subgroups based on AF ablation outcomes and identify diagnostic codes associated with failure.

**Methods:** Machine learning clustering with must-link and cannot-link constraints was applied to electronic health records to discover meaningful clusters, revealing patient-specific factors influencing procedural success or failure. Statistical analyses, including chi-square tests, were used to identify diagnostic codes significantly associated with ablation failure.

**Results:** Out of the 145 diagnostic codes examined, thirteen significant codes were identified and categorized into four primary risk groups, ranked by their impact on procedural outcomes: (1) direct contributors affecting cardiovascular health, (2) indirect factors that contribute to systemic stress, (3) complications related to anticoagulation and hemorrhagic risks that can impact bleeding management, and (4) broader health indicators reflecting a general health burden that reduce patients resilience to procedural stress.

**Conclusions:** This study shows the importance of cardiovascular and non-cardiovascular factors in AF ablation outcomes, emphasizing the need for a more comprehensive pre-procedural evaluation. It also contributes to the application of machine learning in personalized risk assessment for AF and advancing individualized care strategies that may improve ablation success.

## Introduction

Atrial fibrillation (AF) is a common and clinically significant cardiac arrhythmia characterized by rapid and irregular electrical impulses in the atria.^1-3^ This irregularity disrupts normal cardiac function, leading to severe health complications, including stroke, heart failure, cognitive decline, diminished quality of life, and increased mortality.^4-6^ The global burden of AF continues to rise from 59.7 million in 2019, driven by aging populations and the increasing prevalence of comorbidities such as hypertension, diabetes, and obesity.^7-9^ In the United States alone, an estimated 4.48% of the population, approximately 10.55 million adults, were diagnosed with AF in 2019, and this number is projected to exceed 12.1 million by 2030.^10-12^ These trends underscore the urgent need for more effective and personalized management strategies.

Over time, the natural course of AF leads to patients experiencing progressively more frequent and longer-lasting episodes, which can occur sporadically and unpredictably.^13, 14^ Managing AF is further complicated by the need for lifelong anticoagulation to reduce the risk of stroke.^15^. Various treatments are available for AF, and recent research findings from the Early Rhythm-Control Therapy (EAST)^16, 17^ demonstrates the benefits of treatments, such as AF ablation to restore normal sinus rhythm (NSR) early after AF diagnosis. Therefore, AF ablation is increasingly recommended as a first-line therapy to restore NSR for symptomatic paroxysmal or persistent AF.^3, 10, 16, 18^ It is often preferred by patients due to its potential to provide a long-term solution. Despite its widespread use, AF ablation success rates vary considerably, with some patients achieving long-term success while others experiencing recurrent AF or procedural failure.^17, 18^

Sociodemographic and clinical factors have traditionally been used to predict AF ablation outcomes, yet their collective predictive power remains limited.^19^ In this era of striving for targeted patient treatments to improve outcomes, more emphasis should be placed on personalized risk prediction to refine patient selection. However, existing approaches tend to overlook heterogeneity among patients, potentially masking useful insights into why some individuals respond differently to the same treatment. A growing area of research focuses on the use of data-driven approaches to identify patterns and subgroups within patient populations.^20^ Electronic health records (EHRs), which contain comprehensive longitudinal health data, provide a valuable resource for analyzing large patient cohorts. The broad adoption of EHR systems presents a crucial opportunity for transformative change in predicting AF ablation outcomes and offers a new strategy for identifying key predictors of procedural success or failure. A deeper understanding of patient-specific factors in predicting AF ablation outcomes will help refine and personalize patient selection for the procedure.^21^

Recent advances in machine learning (ML), such as cluster analysis, have shown promise in identifying patient subgroups based on shared clinical features.^22, 23^ These techniques, when applied to EHR data, could provide new insights into predicting AF ablation outcomes and developing more personalized therapeutic strategies.

In this study, we explore whether cluster analysis can identify useful subgroups of patients based on their post-ablation outcome trajectories. We aim to assess how these subgroups correlate with both clinical and demographic variables and determine whether such classifications can help inform future treatment decisions and improve patient care.

## Materials and Methods

### Dataset

The MarketScan® research database was used for this study which focuses on claims from individuals enrolled in Medicare Supplemental and Medicare Advantage plans. The study cohort includes 14,610 patients who underwent de novo AF ablation between January 1, 2013, and December 31, 2020. The procedure was identified using CPT code 93656. Each patient had a unique ID and received inpatient admission (I), outpatient service (O), and inpatient service (S) care. AF was confirmed using ICD-9 code 427.31 or ICD-10 code I48.X, ensuring the accurate identification of diagnoses and procedures. These claims data provided insights into patients’ medical histories, particularly focusing on their pre- and post-ablation outcomes. The MarketScan® database contains other claims datasets including Commercial claims, Multi-State Medicaid data, Health and Productivity Management, and more.^24^ However, for this research, we chose the Medicare claims over the others because it primarily include an older population, which is more representative of patients with AF. Additionally, Medicare data provides comprehensive, long-term follow-up, and consistent coverage, offering a more relevant and unbiased dataset for evaluating AF ablation outcomes.^25^

### Preprocessing and feature selection

To ensure consistency and relevance for the post-ablation outcome analysis, the dataset was pre-processed. Firstly, patient records focusing on diagnoses that occurred within one year after AF ablation were extracted, while standardizing both diagnosis and procedure dates. We then captured the frequency of occurrence of diagnosis codes for each patient, providing an overview of the most prevalent medical condition during this critical time frame while minimizing the impact of rare diagnoses. To address Medicare’s transition from ICD-9 to ICD-10 coding after 2015, the ICD-10 codes were converted back to ICD-9 using a lookup tool^26^ ensuring that all diagnosis codes were represented accurately. Furthermore, to keep the dataset streamlined and manageable, we retained only the first three digits of the converted ICD codes (for example, 427.31 becomes 427). Certain codes, such as those starting with E, V, or Z, which typically refer to external factors were excluded from the analysis since they were not directly relevant to the study’s goals.

### Methods

The diagnosis codes including demographic details from the AF patient’s population who had undergone ablation procedures were encoded as binary variables, where a value of 1 indicated the presence of a particular diagnosis, and 0 indicated its absence. Before clustering, non-essential columns such as patient ID and procedure dates were excluded to focus on relevant clinical features.

### Clustering Analysis

We employed the COP-KMeans algorithm, a variant of KMeans that integrates cannot-link constraints.^27-29^ This allowed us to incorporate domain knowledge by ensuring that patients with different ablation outcomes (failure or success) were not assigned to the same cluster. The clustering process started by randomly initializing the centroids of the clusters, selected based on domain understanding of the population. The algorithm iteratively assigned data points to the nearest centroid, respecting the cannot-link constraints, and updated the centroids until convergence was achieved. The clustering performance was evaluated using the silhouette score,^30^ adjusted rand index (ARI),^31^ and mutual information (MI).^32^ The silhouette score reflects how well each data point fits within its assigned cluster compared to other clusters, providing insight into the cohesion and separation of clusters. The ARI evaluates how closely the predicted clusters align with the true cluster labels, accounting for random chance. Meanwhile, MI measures the overlap in information between the predicted clusters and the true labels, showing how well one clustering explains the other.

### Statistical Analysis

Following the clustering, we analyzed the distribution of success and failure outcomes within each cluster. Specifically, we focused on calculating the proportion of failure cases in each cluster. To further investigate the differences between clusters, we conducted pairwise comparisons of diagnosis codes across clusters associated with ablation failure. This analysis was aimed at identifying diagnosis codes that were significantly different between clusters, which could suggest potential subgroups of interest.

### Pairwise Comparison of Clusters

We conducted pairwise comparisons of diagnosis code distributions between clusters with failed outcomes. For instance, in the West region, the failed clusters 0, 3, and 4 in this case were compared using chi-square tests. For each diagnosis code, we computed the mean occurrence within each cluster and used chi-square tests to assess the significance of differences between clusters, identifying diagnosis codes that were more prevalent in certain failure clusters and, providing insights into possible subgroups with distinct clinical profiles.

### Chi-Square Test for Diagnosis Code Significance

To further explore the association between diagnosis codes and ablation outcomes, we performed chi-square tests on the entire dataset. For each diagnosis code, we constructed contingency tables comparing the presence or absence of the code with the ablation outcome (success or failure). This allowed us to compute both the chi-square statistic and the corresponding p-value for each diagnosis code. Additionally, we calculated the proportion of failure cases associated with each code, enabling us to identify diagnosis codes that were most strongly linked to failure outcomes. Diagnosis codes with p-values less than 0.05 and failure proportions greater than or equal to 0.55 were considered significant and were ranked by their failure proportions.

### Ethical considerations

For this study, the data contained no personal health-identifying information. Therefore, the Emory Institutional Review Board (IRB) deemed the study exempt from needing IRB approval.

## Results

### Demographic and clinical characteristics

This analysis included a total of 14,610 patients who underwent AF ablation between January 1, 2013, and December 31, 2020. The mean age of patients was 71.5 years (SD = 5.31), with female patients comprising approximately 40% of the population. About 24.73% of the patients had an additional diagnosis of atrial flutter, often linked to AF and potentially affecting ablation outcomes. Due to the limitations of ICD-9 coding before 2015, categorizing AF subtypes as either persistent or paroxysmal AF was only possible for a subset of 6,981 patients because they had ICD-10 codes. Among these, 59.60% were diagnosed with persistent AF, while 40.40% had paroxysmal AF. Success rates for ablation varied between these groups, with a 59.75% success rate for patients with persistent AF and 40.25% for those with paroxysmal AF. Further demographic and clinical details are outlined in Table 1.

**Table 1.** Demographic and clinical characteristics of patients.

| **Demographic and Clinical Variable** | **Overall (N=14610)** | **AF Ablation Success (N=8141)** | **AF Ablation Failure (N=6469)** |
| --- | --- | --- | --- |
| **Age (mean ± standard deviation)** | 71.6 ± 5.32 | 71.6 ± 5.29 | 71.6 ± 5.34 |
| **Female** | 5839 (39.97%) | 3236 (39.75%) | 2603 (40.24%) |
| **Charlson Comorbidity Index** |  |  |  |
| 0 | 7160 (49.01%) | 3996 (49.08%) | 3164 (48.91%) |
| 1 | 4423 (30.27%) | 2480 (30.46%) | 1943 (30.04%) |
| $\geq 2$ | 3027 (20.72%) | 1665 (20.45%) | 1362 (21.05%) |
| **Elixhauser Comorbidity Index** |  |  |  |
| 0 | 1812 (12.40%) | 1042 (12.80%) | 770 (11.90%) |
| 1 | 4435 (30.36%) | 2474 (30.39%) | 1961 (30.31%) |
| $\geq 2$ | 8363 (57.24%) | 4625 (56.81%) | 3738 (57.78%) |
| Patients with ICD-10 | N=6981 | N=3866 | N=3115 |
| Paroxysmal AF | 2820 (40.40%) | 1556 (40.25%) | 1264 (40.58%) |
| Persistent AF | 4161 (59.60%) | 2310 (59.75%) | 1851 (59.42%) |

### Patient clustering and evaluation

We used a constrained clustering method, specifically COP-KMeans^29, 33^ to categorize patients’ records into distinct subgroups based on the outcomes of their AF ablation procedures. To ensure meaningful groupings, the clustering was based on cannot-link constraints which prevent patients with successful outcomes from being clustered with those who experienced failure. This approach enables us to focus on identifying unique characteristics and patterns among patients with failure outcomes. Multiple evaluation metrics including silhouette score, ARI, and MI were employed to assess the clustering quality. The results indicated moderate performance with a silhouette score of 0.3, an ARI of 0.61, and an MI of 0.68 suggesting that the clustering approach was reasonably effective in grouping patients and capturing meaningful distinctions between subgroups. For further evaluation, t-SNE was used to visualize the clusters which provided additional insight by revealing diverse boundaries in some areas. However, there was some overlap, pointing to commonalities among certain patient subgroups.

### Identification of significant diagnosis and failure proportions

The study analyzed 145 diagnostic codes from the patient records and identified thirteen codes that showed significant associations with AF ablation failure. Using chi-square tests, we examined differences in the prevalence of these codes between patients with successful and unsuccessful ablation outcomes. The results point to specific conditions that may either directly contribute to or indirectly influence the likelihood of ablation failure.

Among the diagnostic codes with the highest failure proportions, we found:

- 569 (Other disorders of intestine) - failure proportion: 58.3%
- 783 (Symptoms concerning nutrition, metabolism, and development) - failure proportion: 57.5%
- 404 (Hypertensive heart and chronic kidney disease) - failure proportion: 57.2%
- 492 (Emphysema) - failure proportion: 56.7%
- 403 (Hypertensive chronic kidney disease) - failure proportion: 56.2%

These findings suggest that some of these conditions may directly impact the risk of ablation failure. In contrast, others may play more of an indirect or contributing role, necessitating closer attention. Table 2 presents the significant diagnosis codes.

**Table 2.** Key co-morbidity indicators of AF ablation failures.

| **Diagnosis Code** | **Code Description** | **Chi-Square** | **P-Value** | **Failure Proportion (%)** |
| --- | --- | --- | --- | --- |
| 404 | Hypertensive heart disease | 26.51 | 2.62e-07 | 57.2 |
| 492 | Emphysema | 19.04 | 1.28e-05 | 56.7 |
| 403 | Chronic kidney disease | 48.14 | 3.97e-12 | 56.2 |
| 411 | Ischemic heart disease | 16.58 | 4.67e-05 | 56.0 |
| 783 | Symptoms concerning nutrition, metabolism, and development | 25.62 | 4.15e-07 | 57.5 |
| 274 | Gout | 28.47 | 9.49e-08 | 55.4 |
| 578 | Gastrointestinal hemorrhage | 24.49 | 7.46e-07 | 55.4 |
| 286 | Coagulation defects | 17.66 | 2.64e-05 | 55.2 |
| 569 | Other disorders of intestine | 39.65 | 3.03e-10 | 58.3 |
| 791 | Symptoms involving urinary system | 16.11 | 5.99e-05 | 55.8 |
| 553 | Hernia of abdominal cavity | 24.52 | 7.34e-07 | 55.3 |
| 211 | Benign neoplasm of other parts of the digestive system | 45.03 | 1.94e-11 | 55.2 |
| 595 | Cystitis | 15.74 | 7.26e-05 | 55.1 |

## Discussion

Clustering algorithm and chi-square test helped identify diagnosis codes that may be contributing to AF ablation failure. This study highlights a range of conditions, both directly and indirectly related to AF ablation outcomes. We have grouped these findings into main categories to better understand their potential impact on ablation failure.

### Direct contributors of failure

Certain codes, such as 404 (hypertensive heart disease), 403 (chronic kidney disease), and 411 (ischemic heart disease) are directly connected to cardiovascular health and likely have a significant impact on ablation outcomes. These conditions are known to strain the heart and complicate blood flow, making successful ablation more challenging. Code 492 (Emphysema) represents a chronic respiratory condition that also affects cardiovascular function and thus may play a role in influencing failure rates. These diagnoses are some of the most prominent direct factors likely contributing to procedural challenges.

### Indirect or contributing factors

Diagnosis codes like 783 (Symptoms concerning nutrition, metabolism, and development) and 274 (Gout) may not directly affect AF outcomes but can indirectly increase risks due to systemic inflammation, metabolic instability, or other underlying health complications. For example, gout is associated with systemic inflammation, which can worsen cardiac stress and impact the recovery process. Similarly, metabolic symptoms associated with code 783 may lead to imbalances that affect overall cardiovascular health.

### Complications in anticoagulation and hemorrhagic risk

Codes related to coagulation defects (286) and gastrointestinal hemorrhage (578) highlight potential complications in anticoagulation management, an essential aspect of AF patient care. Patients with coagulation defects face increased bleeding risks, which can complicate both the procedure and post-procedural recovery, impacting ablation outcomes.

### General Health Profile Indicators

Some of the identified codes, such as 569 (Other disorders of intestine), 791 (Symptoms involving urinary system), 211 (Benign neoplasm of other parts of the digestive system), 553 (Hernia of abdominal cavity), and 595 (Cystitis) may not directly contribute to AF ablation failure but provide a broader view of the health challenges these patients face. While these conditions do not typically impact AF ablation outcomes directly, they reflect a more complex health profile that could reduce a patient’s resilience to procedural stress and therefore affect recovery.

In summary, our analysis highlights that both cardiovascular and non-cardiovascular conditions can influence the risk of AF ablation failure. Direct contributors, such as hypertensive heart disease and chronic kidney disease, pose clear and immediate risks, while metabolic issues, coagulation challenges, and broader health conditions act as indirect but meaningful factors. By categorizing these diagnostic codes into four risk groups, we provide a framework that enables healthcare providers to identify high-risk patients more effectively, tailor assessments, and optimize care strategies. These findings have the potential to improve patient outcomes and advance personalized approaches to managing AF ablation procedures.

## Data Availability

All data produced in the present study are available upon reasonable request to the authors

## Study Limitations

This study relied on electronic health records (EHR) from individuals with Medicare Advantage or Medicare Supplemental plans. Like other administrative records, these may contain some inaccuracies or inconsistencies due to human error. To minimize these concerns, we clearly outlined the data source, inclusion, and exclusion criteria. The analysis also focused on a subset of 145 diagnostic codes, which might not cover all factors influencing ablation outcomes. Finally, while statistical tests were used to identify significant associations, there may still be residual confounding factors influencing the observed relationships between diagnostic codes and ablation outcomes.

## Perspectives

### Competency in Medical Knowledge

AF is a common heart rhythm disorder with success rates for ablation that vary widely. Understanding how specific health conditions contribute to ablation failure can help healthcare professionals assess patient risks better.

### Translational Outlook

The combination of machine learning and statistical analysis to identify risk factors can enable more individualized patient evaluations before ablation, enhancing procedural outcomes and extending the benefits of ablation to a broader range of patients.

## Abbreviations

AF: Atrial Fibrillation
NSR: Normal Sinus Rhythm
EHR: Electronic health records
ML: Machine Learning
SD: Standard Deviation
CPT: Current Procedural Terminology
ICD: International Classification of Disease
ARI: Adjusted Rand Index

